# Does genetic predisposition modify the effect of lifestyle-related factors on DNA methylation?

**DOI:** 10.1101/2021.10.18.21265181

**Authors:** Chenglong Yu, Allison M Hodge, Ee Ming Wong, Jihoon E Joo, Enes Makalic, Daniel F Schmidt, Daniel D Buchanan, Gianluca Severi, John L Hopper, Dallas R English, Graham G Giles, Roger L Milne, Melissa C Southey, Pierre-Antoine Dugué

## Abstract

Lifestyle-related phenotypes have been shown to be heritable and associated with DNA methylation. We aimed to investigate whether genetic predisposition to tobacco smoking, alcohol consumption and higher body mass index (BMI) moderates the effect of these phenotypes on blood DNA methylation. We calculated polygenic scores (PGS) to quantify genetic predisposition to these phenotypes using training (*N*=7,431) and validation (*N*=4,307) samples. Using paired genetic-methylation data (*N*=4,307), gene-environment interactions (i.e. PGS x lifestyle) were assessed using linear mixed-effects models with outcomes: 1) methylation at sites found to be strongly associated with smoking (1,061 CpGs), alcohol consumption (459 CpGs) and BMI (85 CpGs), and 2) two epigenetic aging measures, *PhenoAge* and *GrimAge*. In the validation sample, PGS explained ∼1.4% (P=1×10^−14^), ∼0.6% (P=2×10^−7^) and ∼8.7% (P=7×10^−87^) of variance in smoking initiation, alcohol consumption and BMI, respectively. Nominally significant interaction effects (P<0.05) were found at 61, 14, and 7 CpGs for smoking, alcohol consumption and BMI, respectively. There was strong evidence that all lifestyle-related phenotypes were positively associated with *PhenoAge* and *GrimAge*, except for alcohol consumption with *PhenoAge*. There was weak evidence that the association of smoking with *GrimAge* was attenuated in participants genetically predisposed to smoke (interaction term: -0.02, P=0.06) and that the association of alcohol consumption with *PhenoAge* was attenuated in those genetically predisposed to drink alcohol (interaction term: -0.03, P=0.04). In conclusion, genetic susceptibility to unhealthy lifestyles did not strongly modify their effects on blood DNA methylation. Potential associations were observed for epigenetic aging measures, which should be replicated in additional studies.

## Introduction

Strong evidence shows that lifestyle-related factors influence epigenetic regulation mechanisms, such as DNA methylation [1-3]. For instance, tobacco use [4], alcohol consumption [5] and excess body weight [6] have been found to be strongly associated with blood DNA methylation changes. These lifestyle-related phenotypes are to some extent heritable [7-11]. Meta-analysis studies using imputed genotype data have estimated the genetic predisposition of an individual to smoke, drink alcohol or become overweight or obese, with SNP-based heritability estimates of ∼8% for smoking initiation [7], ∼4% for alcohol drinks per week [7] and ∼22% for body mass index (BMI) [8]. Therefore, a key question of interest is whether such genetic predisposition to unhealthy lifestyle-related factors modifies their relationship with DNA methylation.

One way to understand such modification lies in examining gene-environment (i.e. gene-lifestyle) interactions in DNA methylation. Many gene-lifestyle interaction studies have focused on single genetic variants in candidate genes using Asian-ancestry samples [12-17]. For instance, strong interactions between the effects of alcohol intake and smoking on oesophageal and gastric cancer with risk alleles in alcohol dehydrogenase (*ADH*) and aldehyde dehydrogenase (*ALDH*) genes have been found in large-scale population-based Japanese studies [12-14]. However, Ugai *et al*. found no evidence of a gene-environment interaction between the *ALDH2* alcohol-consumption predisposition variant (rs671) and alcohol intake for breast cancer risk among Asian women from the Breast Cancer Association Consortium [17]. Thus, individual genetic variants, which have small effect sizes in association with complex lifestyle-related traits, may not always be informative for evaluating overall genetic susceptibility. Furthermore, these variants are typically very rare among European-ancestry populations compared to Asian-ancestry groups. For example, the minor allele frequency (MAF) of *ALDH2* rs671 is 0% in TwinsUK registry, while it is 16%, 19% and 21% for the Korean, Japanese and Vietnamese populations, respectively [18]. Therefore, when using single variants, lack of power may limit ability to detect interactions, especially for European-ancestry population-based data.

In contrast to candidate genes, polygenic scores (PGS), which summarise the estimated effects of many variants into a single value, have emerged as a powerful way to quantify an individual’s genetic predisposition to a phenotype [19]. In previous studies, PGS were found to account for approximately 4%, 2.5% and 14% of variance in smoking initiation [7], alcohol consumption per week [7] and BMI [8], respectively, i.e. about half of the estimated SNP-based heritability of these phenotypes.

In this study, we hypothesised that genetic factors that confer a predisposition to unhealthy lifestyle-related phenotypes would moderate their harmful effects - for example, individuals genetically predisposed not to drink would likely drink less over their lifetime but might have a higher-than-average risk of associated disease. We used DNA methylation as an outcome variable in the analyses since many diseases have been shown to be associated with aberrant DNA methylation [20-26], including measures of epigenetic aging [27-31]. Our aim in this study was therefore to investigate the interaction effects of observed tobacco smoking, alcohol consumption, and BMI with their respective PGS on DNA methylation in blood, for i) loci at which methylation changes with lifestyle and ii) two measures of epigenetic aging, *PhenoAge* and *GrimAge*.

## Materials and Methods

### Study participants

The Melbourne Collaborative Cohort Study (MCCS) is an Australian community-based study that recruited 41,513 European-ancestry participants in 1990-1994 [32]. Several nested case-control studies have been conducted to evaluate associations between blood DNA methylation and the risk of eight types of cancer [33-36]. DNA was extracted from pre-diagnostic peripheral blood taken at recruitment (1990-1994) or at a subsequent follow-up visit (2003-2007) in cancer-free participants. Incident cases were matched to controls on age, sex, country of birth and sample type (buffy coats, dried blood spots, and peripheral blood mononuclear cells) using incidence density sampling [32]. We used self-reported questionnaire-collected data on tobacco use, alcohol consumption, and measured height and weight to calculate BMI [4-6] for participants in the MCCS. The study was approved by the Cancer Council Victoria’s Human Research Ethics Committee, Melbourne, VIC, Australia, and all participants provided informed consent in accordance with the Declaration of Helsinki.

### Genetic and DNA methylation data

Genome-wide genotyping was conducted on blood DNA samples from 12,584 MCCS participants using the Infinium OncoArray-500K BeadChip (Illumina, San Diego, CA, USA) [32, 37]. Following previous standardised protocols [38], we imputed autosomal genotypes using the Michigan imputation server [39] and IMPUTE version 2 [40] with the 1000 Genomes Project dataset (phase 3) as the reference panel. The genotype probabilities from imputation were used to hard-call (uncertainty < 0.1) the genotypes for variants with an imputation info score□>□0.3. We then retained the hard-called variants with MAF >□0.1%, missing genotype rate□<□10% and Hardy-Weinberg equilibrium P-value□>□10^−6^. To avoid bias due to confounding by shared environment among close relatives, individuals were removed based on relatedness by excluding one participant randomly selected from any pair with a genetic relationship > 0.125 (3rd-degree or closer relationship) using the software GCTA [41]. After these quality control (QC) steps, 11,942 unrelated individuals with 9,355,361 genetic variants (including 8,578,993 SNPs) were retained for the follow-up analyses.

We measured DNA methylation in blood samples from 4,511 of the 11,942 participants using the HumanMethylation450 BeadChip (Illumina, San Diego, CA, USA) applying methods described previously [32, 42, 43]. Among the 4,511 participants, DNA of 4,307 were extracted at baseline recruitment (1990-1994). QC details for measures of genome-wide DNA methylation have been reported previously [34-36, 44]. Briefly, we removed probes with missing rate > 20% and probes on Y-chromosome, and ultimately retained 484,431 CpG sites with their beta values for each sample. Of these, we focused on 1,061, 459 and 85 CpG sites (**Tables S1-S3**) that were found to be strongly associated with smoking, alcohol consumption and BMI, respectively, with P < 10^−7^ in both the MCCS and external data [4-6, 34]. Methylation M-values, calculated as log_2_(beta/(1-beta)), were used as these are thought to be more statistically valid for detection of differential methylation [45].

The 7,431 individuals with genetic data but without DNA methylation data were used as the PGS training sample, and the 4,307 individuals with paired genetic-methylation data were used for PGS validation and all other analyses of this study (**Table 1**).

**Table 1.** Characteristics of the MCCS participants used in the study.

| Sample characteristic | Training sample for PGS<br>(*N* = 7,431) | Validation sample for PGS and for main analysis (*N* = 4,307) |
| --- | --- | --- |
| Age at blood draw (median [IQR]) | 53.7 [47.1 - 60.8] | 59.6 [52.7 - 64.8] |
| Sex: |  |  |
| male, *N* (%) | 2,655 (36%) | 2,541 (59%) |
| female, *N* (%) | 4,776 (64%) | 1,766 (41%) |
| Blood sample type: |  |  |
| dried blood spots, *N* (%) |  | 3,240 (75%) |
| peripheral blood mononuclear cells, *N* (%) |  | 993 (23%) |
| buffy coats, *N* (%) |  | 74 (2%) |
| Smoking status: |  |  |
| current, *N* (%) | 644 (9%) | 477 (11%) |
| former, *N* (%) | 2,235 (30%) | 1,638 (38%) |
| never, *N* (%) | 4,552 (61%) | 2,192 (51%) |
| Alcohol consumption last week (g/day) *, median [IQR] | 2.7 [0 – 14.7] | 4.3 [0-17.1] |
| Body mass index (kg/m<sup>2</sup>), median [IQR] | 26.1 [23.6 - 28.9] | 26.6 [24.3 - 29.4] |

\*Note: Participants reported their alcohol intake on each day during the previous week, in terms of the number, measure, and type of drink (beer, wine, spirits). Grams per day (g/day) were then calculated based on average number of drinks a participant reported, as described previously [5, 41].

### PGS analyses

We considered three lifestyle-related phenotypes: tobacco use, alcohol consumption and BMI. The largest published GWAS to date for smoking and alcohol consumption (∼1,200,000 samples) [7] and for BMI (∼700,000 samples) [8] were used as base data, which provided estimated effect and P-value for each genetic variant; we used the same phenotypic definitions and variable transformations as those used in these GWAS [7, 8].

For tobacco use, we used smoking initiation [7] - a dichotomous phenotype for participants reporting ever being a regular smoker in their lifetime (current or former smokers), and those who reported never being a regular smoker. For alcohol consumption, participants reported their alcohol intake on each day during the previous week, in terms of the quantity and type of drink (beer, wine, spirits). Grams per day (g/day) were calculated based on the average number of drinks a participant reported, as described previously [5, 46]. The variable was left-anchored at 1 and log-transformed to minimise the influence of potential outliers [7]. For BMI, we applied a rank-based inverse normal transformation to the raw values (in kg/m^2^) to better approximate a normal distribution [8].

PGS were calculated using the PRSice software [47, 48] with LD clumping parameters of R^2^ > 0.25 over 250-kb sliding windows. We removed ambiguous SNPs with A/T or G/C alleles [48]. A total of 7,431 MCCS individuals were used as a training dataset (“target data” in PRSice), and the PGS with the P-value threshold (from 5×10^−8^ to 1 by increments of 5×10^−5^) found to explain most of the variance in the phenotypes in the training dataset was chosen as the optimal polygenic score. The phenotypic values were adjusted for age, sex and first 20 ancestry principal components (PCs) to account for population structure. To assess if there was overfitting in the association of optimal PGS with phenotype [48], we used the 4,307 MCCS individuals, which were unrelated to the training dataset, as an out-of-sample validation.

For comparison with the optimal PGS, and to retain only SNPs having a strong effect on phenotypes of interest, we also calculated a ‘genome-wide significant PGS’ using only variants with a P-value < 5×10^−8^ and LD clumping parameters of R^2^ > 0.25 over 250-kb sliding windows, and evaluated the association of this PGS with phenotype for both training and validation datasets.

### Statistical analyses

Using the 4,307 MCCS participants with paired genetic-methylation data, we examined the interaction effects of observed tobacco use, alcohol consumption, and BMI with their respective PGS on DNA methylation at individual CpG sites (1,061, 85, and 459 CpGs associated with smoking, alcohol consumption and BMI, respectively [4-6, 34]), using linear mixed-effects regression models with M-values as an outcome. The model was also adjusted for age, sex, first 20 ancestry PCs, sample type, white blood cell composition (percentage of CD4+ T cells, CD8+ T cells, B cells, NK cells, monocytes and granulocytes estimated using the Houseman algorithm [49]) and the two other lifestyle-related phenotypes as fixed effects, and study, batch plate, and chip as random effects. Interaction effects were then assessed by examining the interaction term using the Wald test. It is noted that in these association analyses, we used a continuous comprehensive smoking index (CSI) variable [4] as it includes more information about smoking (cumulative lifetime exposure to tobacco smoke) than smoking initiation, which does not distinguish between current and former smokers or consider other smoking-related variables. For alcohol consumption and BMI, we used the phenotypic values defined in section “PGS analyses”.

We also used the same models to examine interaction effects of these lifestyle-related phenotypes with their PGS on epigenetic aging measures, namely *PhenoAge* [27] and *GrimAge* [28], two composite predictors of mortality. These were calculated using the online calculator [50], and adjusted for age as described previously [31].

## Results

Sample characteristics of the MCCS participants used in this study are shown in **Table 1**.

We calculated optimal and genome-wide significant PGS of three lifestyle-related phenotypes for each participant, summarised in **Table 2**. In the validation sample, we found that the optimal PGS (derived from a training sample, see **Figures S1-S3**) explained ∼1.4% (P = 1×10^−14^), ∼0.6% (P = 2×10^−7^) and ∼8.7% (P = 7×10^−87^) of variance in smoking initiation, alcohol consumption in previous week and BMI, respectively. The genome-wide significant PGS included substantially fewer SNPs and explained less variance.

**Table 2.** PGS calculation for three lifestyle-related phenotypes in training and validation sample

| Lifestyle-related phenotype | **Training sample (N=7,431)** |  |  |  |  |  |
| --- | --- | --- | --- | --- | --- | --- |
|  | Optimal PGS |  |  | Genome-wide significant PGS |  |  |
|  | SNP number | R<sup>2</sup> | P | SNP number | R<sup>2</sup> | P |
| Smoking initiation | 39,813 | 0.014 | 1.4x10<sup>-24</sup> | 145 | 0.004 | 2.9x10<sup>-07</sup> |
| Alcohol consumption in previous week | 151,167 | 0.006 | 5.3x10<sup>-12</sup> | 79 | 0.002 | 4.8x10<sup>-4</sup> |
| BMI | 156,400 | 0.085 | 2.4x10<sup>-146</sup> | 1,877 | 0.041 | 8.5x10<sup>-70</sup> |
| Lifestyle-related phenotype | **Validation sample (N=4,307)** |  |  |  |  |  |
|  | Optimal PGS |  |  | Genome-wide significant PGS |  |  |
|  | SNP number | R<sup>2</sup> | P | SNP number | R<sup>2</sup> | P |
| Smoking initiation | 39,813 | 0.014 | 1.5x10<sup>-14</sup> | 145 | 0.002 | 0.009 |
| Alcohol consumption in previous week | 151,167 | 0.006 | 2.0x10<sup>-7</sup> | 79 | 0.005 | 1.4x10<sup>-6</sup> |
| BMI | 156,400 | 0.087 | 6.6x10<sup>-87</sup> | 1,877 | 0.039 | 3.9x10<sup>-39</sup> |

Note: The SNPs included in optimal PGS were obtained using a P-value threshold (in base data, i.e. external GWAS) that maximised the variance explained in the phenotypes in the training sample, while the SNPs in ‘genome-wide significant PGS’ were obtained using a P-value threshold of $5 \times 10^{-8}$ . R<sup>2</sup> and P are the variance explained and strength of association between the phenotype and its PGS. The validation sample used the same SNPs as those retained in the optimal and genome-wide significant PGS from the training process.

The interactions between the three lifestyle-related phenotypes and their respective optimal PGS in association with DNA methylation at 1,061 smoking-associated CpGs, 459 alcohol consumption-associated CpGs and 85 BMI-associated CpGs are shown in **Tables S1-S3**. Considering a nominal significance threshold of P < 0.05, the numbers of CpGs with significant interaction effects are shown in **Table 3**, none of them being greater than expected by chance. Considering a Bonferroni significance threshold for each of the three phenotypes (P < 0.05/1061 = 4.7×10^−5^, P < 0.05/459 = 1.1×10^−4^ and P < 0.05/85 = 5.9×10^−4^, respectively), we found a significant interaction for a BMI-associated CpG, cg11376147, chr11:57261198 (BMI main effect = -0.06, 95% CI: -0.08, -0.03, P = 8.7×10^−7^; interaction effect with PGS = - 0.04, 95% CI: -0.06, -0.02, P = 3.8×10^−4^), **Table S3**.

**Table 3.**
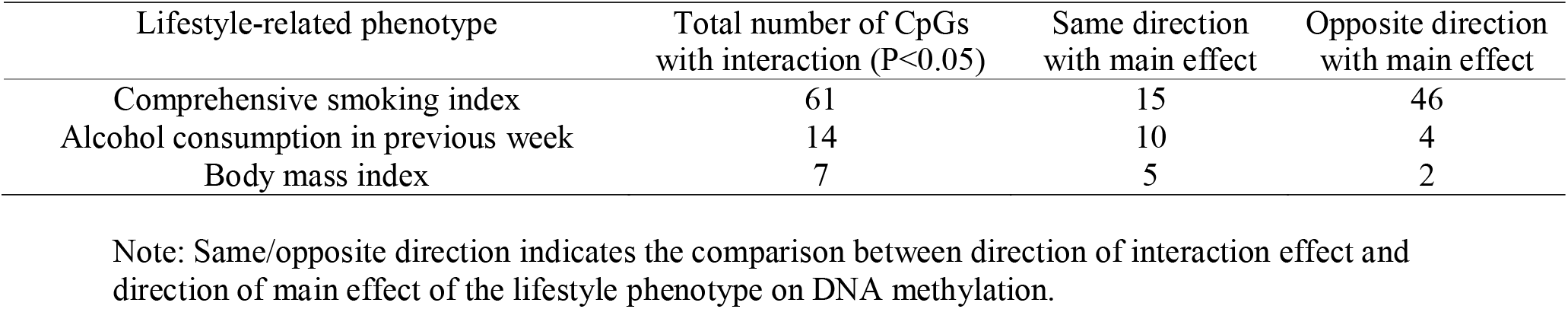
CpG numbers of nominally significant interaction effects between three lifestyle-related phenotypes and their optimal PGS on DNA methylation at 1,061 smoking-associated CpGs, 459 alcohol consumption-associated CpGs and 85 BMI-associated CpGs.

| Lifestyle-related phenotype | Total number of CpGs<br>with interaction (P<0.05) | Same direction<br>with main effect | Opposite direction<br>with main effect |
| --- | --- | --- | --- |
| Comprehensive smoking index | 61 | 15 | 46 |
| Alcohol consumption in previous week | 14 | 10 | 4 |
| Body mass index | 7 | 5 | 2 |

Note: Same/opposite direction indicates the comparison between direction of interaction effect and direction of main effect of the lifestyle phenotype on DNA methylation.

The interactions between the lifestyle-related phenotypes and their optimal PGS in association with *PhenoAge* and *GrimAge* are shown in **Table 4**. There was strong evidence that all lifestyle-related phenotypes were positively associated with *PhenoAge* and *GrimAge*, except for alcohol consumption with *PhenoAge* (P = 0.09). There was weak evidence that the association of CSI with *GrimAge* was attenuated in participants genetically predisposed to smoke with interaction term: -0.022 (95% CI: -0.046, 0.002), P = 0.06 and that the association of alcohol consumption with *PhenoAge* was attenuated in those genetically predisposed to drink alcohol with interaction term: -0.03 (95% CI: -0.059, -0.001), P = 0.04.

**Table 4.** Interaction effects of lifestyle-related phenotypes and their optimal PGS on *PhenoAge* and *GrimAge*.

| Lifestyle-related phenotype | Outcome | Effect (S.E.) of phenotype | P | Effect (S.E.) of PGS | P | Effect (S.E.) of interaction | P |
| --- | --- | --- | --- | --- | --- | --- | --- |
| Comprehensive smoking index | *PhenoAge* | 0.092 (0.015) | $9.2 \times 10^{-10}$ | -0.012 (0.015) | 0.41 | -0.017 (0.014) | 0.22 |
| | *GrimAge* | 0.506 (0.012) | $< 5 \times 10^{-308}$ | 0.030 (0.012) | 0.01 | -0.022 (0.012) | 0.06 |
| Alcohol consumption in previous week | *PhenoAge* | 0.025 (0.014) | 0.09 | -0.016 (0.015) | 0.28 | -0.030 (0.015) | 0.04 |
| | *GrimAge* | 0.072 (0.012) | $1.0 \times 10^{-9}$ | 0.001 (0.012) | 0.91 | 0.018 (0.012) | 0.14 |
| Body mass index | *PhenoAge* | 0.069 (0.015) | $5.7 \times 10^{-6}$ | 0.049 (0.016) | 0.002 | 0.006 (0.014) | 0.68 |
| | *GrimAge* | 0.083 (0.012) | $1.9 \times 10^{-11}$ | 0.013 (0.013) | 0.32 | -0.014 (0.011) | 0.22 |

The results using the genome-wide significant PGS at CpGs of interest are also shown in **Tables S1-S3** and summarized in **Table S4**. Using the Bonferroni correction, a potential interaction was detected at an alcohol-associated CpG cg02470690, chr6:27839548 (alcohol consumption main effect = -0.06, 95% CI: -0.09, -0.03, P = 3.5×10^−5^; interaction effect with PGS = 0.06, 95% CI: 0.03, 0.09, P = 6.7×10^−5^), but not for other lifestyle-related phenotypes or measures of epigenetic aging (**Table S5**).

## Discussion

This is to our knowledge the first study to examine gene-lifestyle interactions on DNA methylation. Interaction studies using PGS are likely to be more powerful than those based on individual variants or genes due to better quantifying the overall genetic predisposition. We expected that genetic predisposition to unhealthy lifestyle-related phenotypes would moderate their harmful effects; however, our results suggest that, for CpGs associated with smoking, alcohol consumption and BMI, genetic predisposition to unhealthy lifestyle-related factors does not strongly modify their effect on blood DNA methylation, i.e. there was no substantial evidence of interaction effects between observed lifestyle-related phenotypes and their respective PGS on DNA methylation at these loci. After Bonferroni correction, an interaction was detected at a BMI-associated CpG (cg11376147); however, the direction of the observed interaction (same as main effect) was inconsistent with our hypothesis. Our results also suggest that the association of smoking with *GrimAge* was slightly attenuated in participants genetically predisposed to smoke and the association of alcohol consumption with *PhenoAge* was slightly attenuated in those genetically predisposed to drink alcohol; the evidence of interaction was nevertheless quite weak and not consistently observed across all lifestyle-related factors and epigenetic aging measures.

In this study, we focused primarily on a best-fit PGS which explains the highest proportion of phenotypic variation, and also considered a PGS including only genome-wide significant variants. Although the latter accounts for less variance, it consists of a number of loci with stronger genetic effects on the lifestyle-related phenotypes, thus this PGS may be more specific to biological mechanisms explaining unhealthy lifestyle and could rule out some confounding effects due to pleiotropy with other traits. For instance, many of the 79 SNPs used for building genome-wide significant PGS of alcohol intake are in *ADH* and *ALDH* genes which are primary enzymes to metabolise alcohol in liver [51], whereas the majority of the 151,167 SNPs included in the optimal PGS may relate to many other traits such as mental health / disorders or educational attainment that are less likely to modify the effects of alcohol consumption on DNA methylation. It is noted that the variance explained (R^2^) in alcohol consumption in the validation sample reduced only a little, from 0.006 to 0.005, when using the optimal and genome-wide significant PGS. This implies that a large number of SNPs in the optimal PGS may not be relevant to this phenotypic trait. Consistent with this, we detected a significant interaction at an alcohol consumption-associated CpG (cg02470690) with the genome-wide significant PGS, with a direction consistent with our hypothesis (opposite to main effect). We also found nominally significant interactions (P < 0.05) that were in the opposite direction to main effect at 41 out of the 459 alcohol-related CpGs, which was substantially more than expected by chance (N=11) and consistent with our previous finding using a PGS of 13 genetic variants associated with alcohol consumption [5].

For alcohol consumption, previous interaction studies were performed predominantly for the *ADH* and/or *ALDH* genes on Asian-ancestry samples [12-17]. In the present study, we used an Australian cohort of European-ancestry, in which the allele frequencies of variants in these alcohol-metabolism genes were very low, most of them were < 1%. For instance, the *ALDH2* rs671 polymorphism, which has been widely studied in Asian groups [52-54], was not included in our analyses due to very low MAF (< 0.1%). Therefore, the candidate gene approach focusing on single variants may not be appropriate in European-ancestry populations for this phenotypic trait, unless resources with a very large sample size are used.

There are several limitations in this study. First, the genotyping was conducted using the Infinium OncoArray-500K BeadChip. Although we performed high-quality imputation to obtain a comprehensive genome-wide range of genotypes, the phenotypic variance explained by PGS we observed was somewhat smaller than using more general microarrays, e.g. R^2^ reached ∼10% for BMI using the same PGS calculation method but with an Illumina 610-Quadc1 array in another study [55]. Secondly, in this study we applied the PRSice method (p-value thresholding and clumping) to calculate PGS but other methods such as Bayesian approaches or regularised regression might have led to somewhat different PGS [56], which may have had some influence on our results. Thirdly, although we included >4,300 participants with paired genetic-methylation data, this sample size might not be sufficient to detect associations at individual or aggregated CpG sites, because interaction effects are usually much smaller than main effects.

In conclusion, genetic susceptibility to unhealthy lifestyles (smoking, drinking alcohol, and being overweight or obese) does not appear to strongly modify their effects on blood DNA methylation based on our data. Potential interactions were observed for the composite measures of epigenetic aging, which suggests that such genetic predisposition might moderate their harmful effects on biological aging, but these findings should be replicated in additional studies.

## Data Availability

All data produced in the present study are available upon reasonable request to the authors

## Supplementary material

Supplemental data for this article (3 supplemental figures and 5 supplemental tables) can be accessed online.

