## Supplementary Figures for "Does genetic predisposition modify the effect of lifestyle-related factors on DNA methylation?"

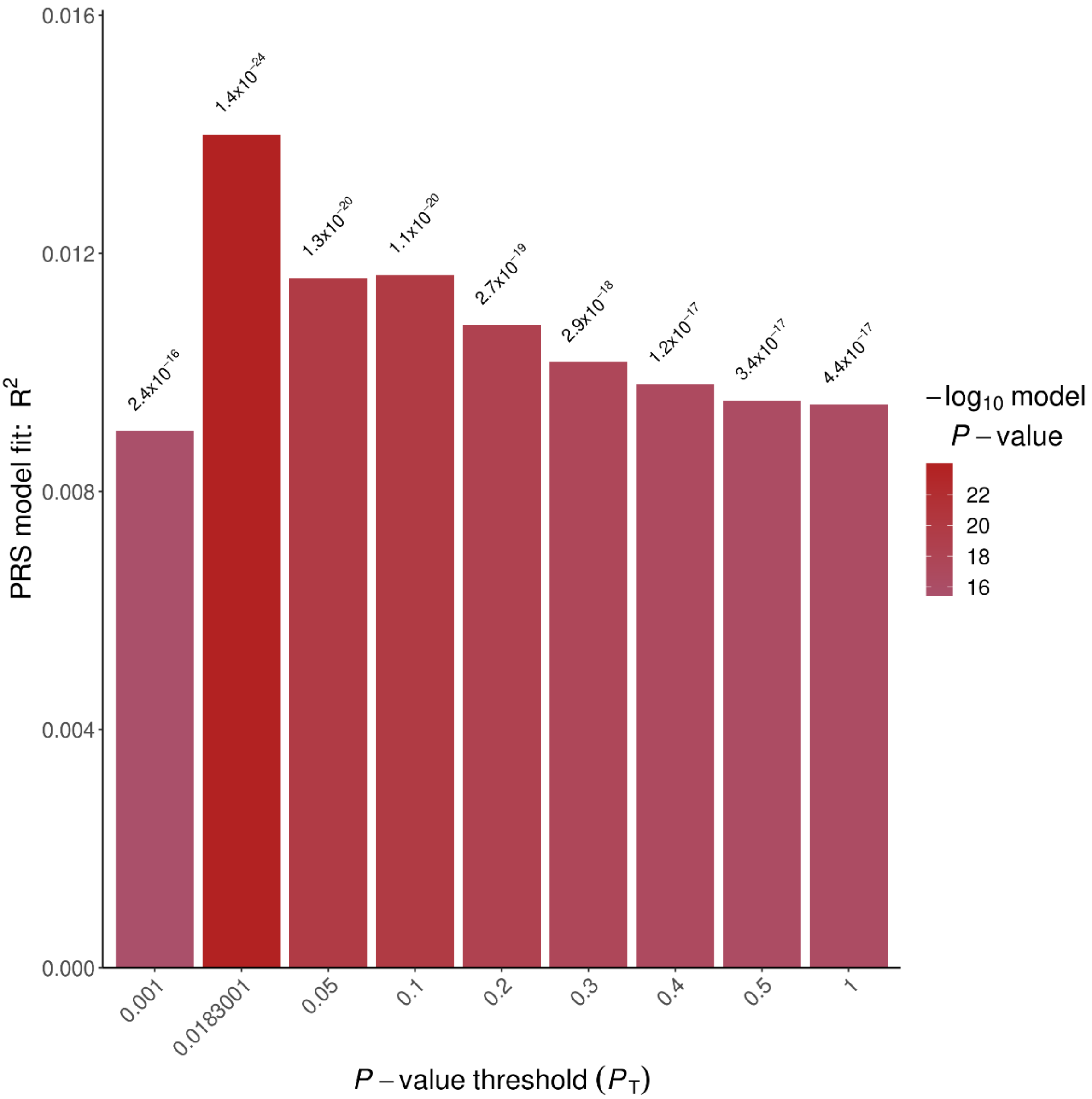


**Figure S1**: Proportion of variance of smoking initiation explained by PGS ($R^2$) in the training sample (*N*=7,431) for different *P*-value thresholds. *P*-value threshold = 0.0183001 generates the best-fit PGS with $R^2$ = 0.014.

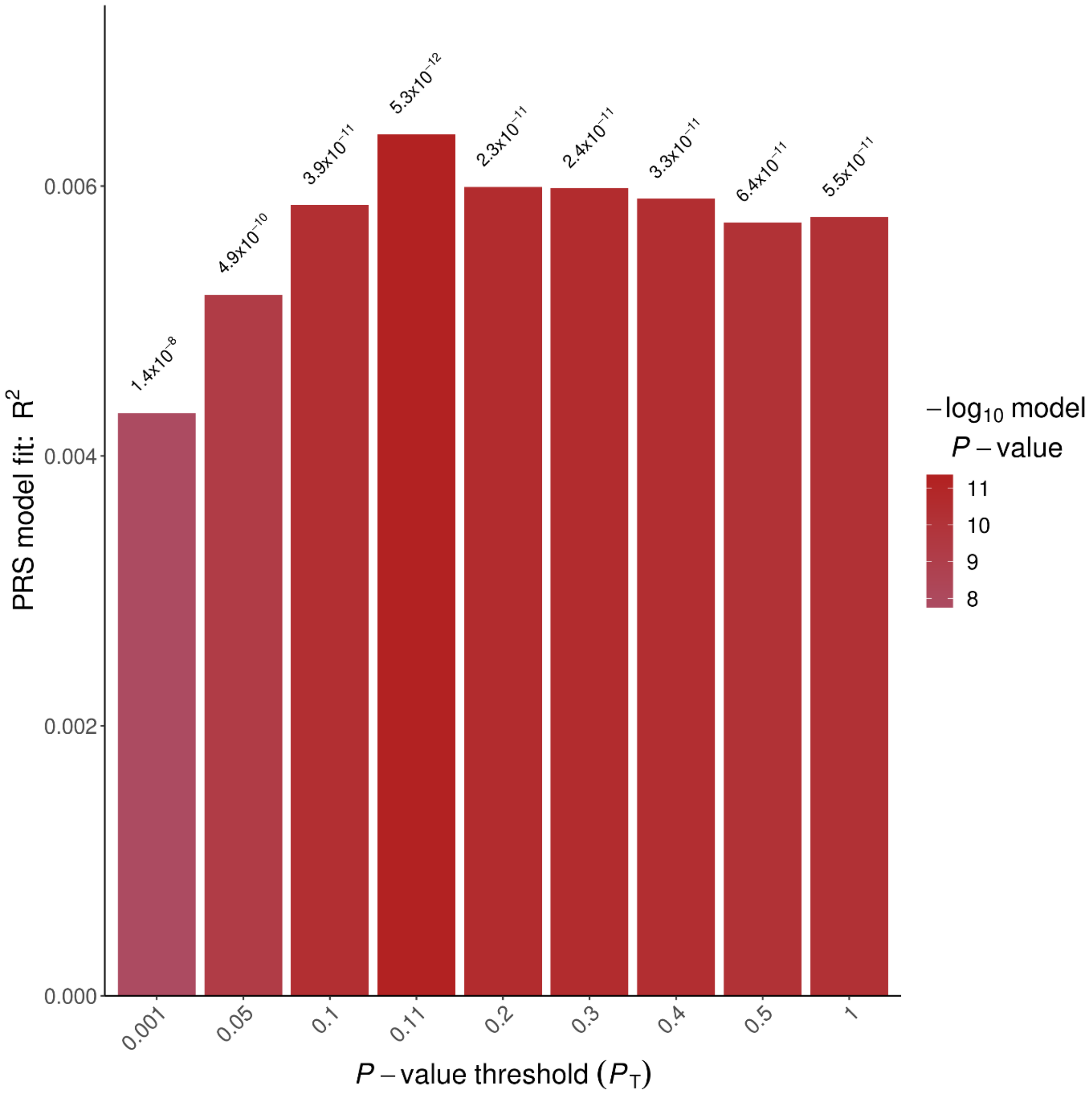


**Figure S2**: Proportion of variance of alcohol consumption last week explained by PGS ($R^2$) in the training sample (*N*=7,431) for different *P*-value thresholds. *P*-value threshold = 0.11 generates the best-fit PGS with $R^2$ = 0.0064.

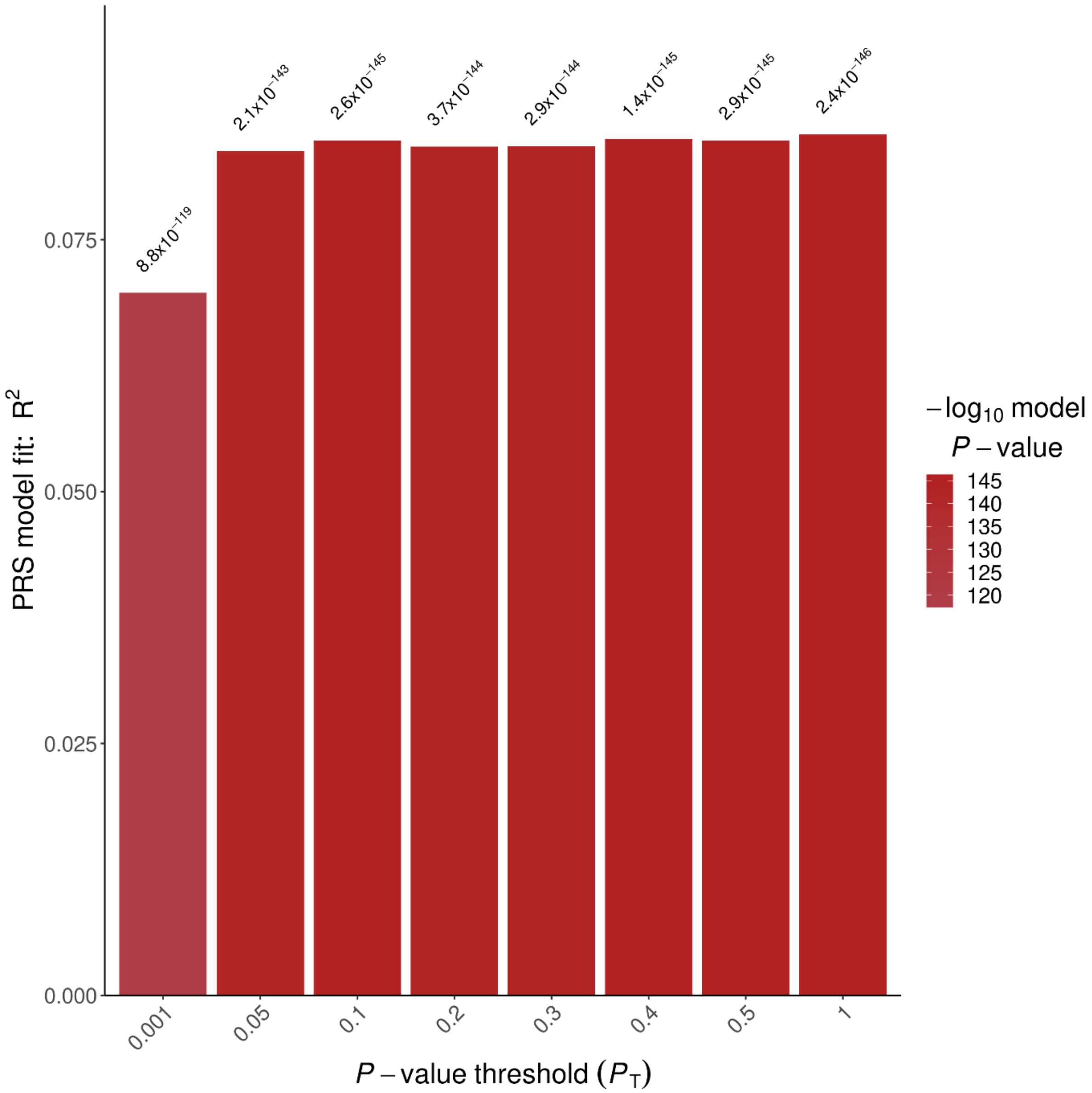


**Figure S3**: Proportion of variance of BMI explained by PGS ($R^2$) in the training sample (*N*=7,431) for different *P*-value thresholds. *P*-value threshold = 1 generates the best-fit PGS with $R^2$ = 0.0855.
